# No evidence of association between *PGK1* variants and Parkinson’s disease

**DOI:** 10.64898/2026.08.18.26360396

**Authors:** Leah V. Chifamba, Sitki Cem Parlar, Lang Liu, Eric Yu, Ziv Gan-Or, Konstantin Senkevich

**Author notes:** **Corresponding author:** Konstantin Senkevich, Montreal Neurological Institute, McGill University, 1033 Pine Avenue, West, Ludmer Pavilion, room 312, Montréal, QC, H3A 1A1.

## Abstract

**Background:** An X-linked levodopa-responsive parkinsonism-epilepsy syndrome has been associated with *PGK1*, and the gene lies within the previously suspected PD locus *PARK12*.

**Objective:** To examine the association of common and rare *PGK1* variants with PD.

**Methods:** We analyzed common and rare variants from Accelerated Medicines Partnership - Parkinson’s Disease (AMP-PD) and UK Biobank (UKBB, total N=4,523 PD cases, 19,736 proxy cases, and 390,532 controls). To account for the X-linked location of *PGK1*, we used sex-stratified, combined regression models and optimized sequence Kernel association (SKAT-O) tests, followed by meta-analysis using MetaSKAT.

**Results:** We found no association between common or rare *PGK1* variants and PD in sex-stratified or combined analyses, including after cross-cohort meta-analysis.

**Conclusion:** Although we did not find evidence supporting an association between *PGK1* and PD, very rare pathogenic *PGK1* variants may still contribute to syndromic parkinsonism. Future research could explore larger datasets to further examine this potential association.

## Introduction

Parkinson’s disease (PD) has a multifactorial pathophysiology, with altered glucose metabolism implicated as a contributing factor (Guimarães et al. 2024). One glycolytic enzyme of interest is phosphoglycerate kinase 1 (PGK1), encoded by the X-linked *PGK1* gene. As the first ATP-generating enzyme in glycolysis, PGK1 plays a vital role in cellular energy metabolism (Zhang et al. 2022; Sakaue et al. 2017). Mutations in *PGK1* result in reduced enzyme activity and impaired cellular energy production, leading to the rare clinical syndrome of PGK1 deficiency (Chiarelli et al. 2012). The disorder exhibits a broad phenotypic spectrum, including chronic hemolytic anemia, myopathy and neurological symptoms such as seizures, stroke-like episodes, and parkinsonism (Morales-Briceño et al. 2019; Sotiriou et al. 2010; Baba et al. 2018).

PGK1 may be relevant to the energy metabolism deficits implicated in PD pathophysiology (Siddique and Kale 2023). Furthermore, *PGK1* lies within the suspected PD locus *PARK12* (Chen et al. 2025; Sakaue et al. 2017). Multiple reports have identified early-onset, levodopa-responsive parkinsonism in individuals with PGK1 deficiency (Guimarães et al. 2024), harboring distinct pathogenic variants, including p.Asp164Val (Morales-Briceño et al. 2019), p.Thr378Pro (Sotiriou et al. 2010; Virmani et al. 2014), p.Ala353Pro (Sakaue et al. 2017) and p.Gly317Asp (Guimarães et al. 2024), highlighting the importance of investigating *PGK1* in PD.

Our study aimed to explore the role of *PGK1* in PD by examining associations of common and rare variants with PD. We analyzed data from two cohorts, comprising 4,523 PD cases, 19,736 proxy cases, and 390,532 controls. The cohorts were first stratified by sex and then combined to account for the X-linked location of *PGK1*.

## Methods

### 2.1 Participants

The population for the genetic analysis consisted of two cohorts, including 4,523 PD patients, 19,736 proxy-patients and 390,532 controls (Table 1). The two cohorts included (i) whole-genome sequencing (WGS) data from Accelerated Medicines Partnership - Parkinson’s Disease (AMP-PD), (https://amp-pd.org/), and it included the Harvard Biomarkers Study, the National Institute of Neurological Disorders and Stroke (NINDS) Parkinson’s Disease Biomarkers Program, the BioFIND study, the NINDS Study of Isradipine as a Disease Modifying Agent in Subjects With Early Parkinson’s Disease (phase 3), and the National Institute on Aging International Lewy Body Dementia Genetics Consortium Genome Sequencing in Lewy Body Dementia case-control cohort.

**Table 1.**
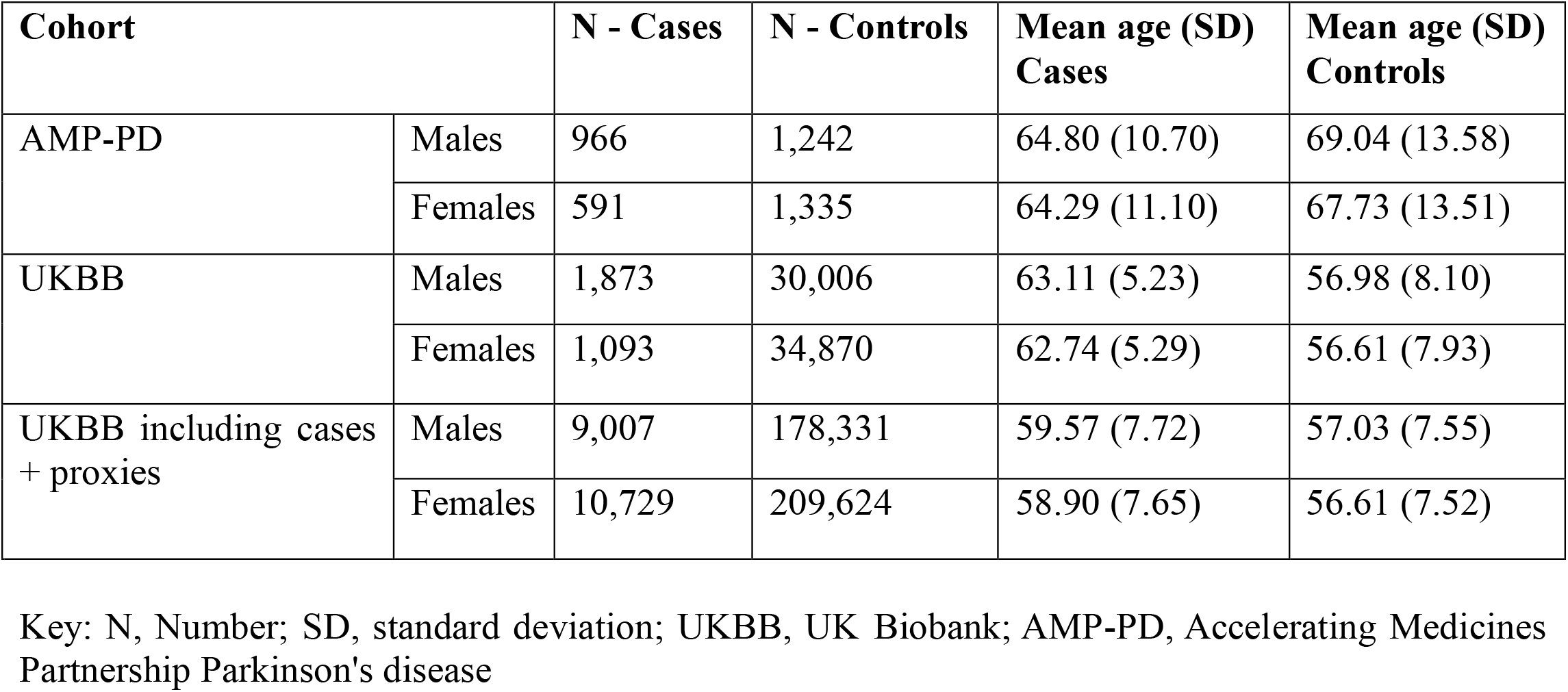
Demographic data of studied cohorts.

| **Cohort** |  | **N - Cases** | **N - Controls** | **Mean age (SD)**<br>**Cases** | **Mean age (SD)**<br>**Controls** |
| --- | --- | --- | --- | --- | --- |
| AMP-PD | Males | 966 | 1,242 | 64.80 (10.70) | 69.04 (13.58) |
|  | Females | 591 | 1,335 | 64.29 (11.10) | 67.73 (13.51) |
| UKBB | Males | 1,873 | 30,006 | 63.11 (5.23) | 56.98 (8.10) |
|  | Females | 1,093 | 34,870 | 62.74 (5.29) | 56.61 (7.93) |
| UKBB including cases<br>+ proxies | Males | 9,007 | 178,331 | 59.57 (7.72) | 57.03 (7.55) |
|  | Females | 10,729 | 209,624 | 58.90 (7.65) | 56.61 (7.52) |

Key: N, Number; SD, standard deviation; UKBB, UK Biobank; AMP-PD, Accelerating Medicines Partnership Parkinson's disease

(ii) The UK Biobank (UKBB) including proxy-cases (first-degree relatives of PD patients) was also acquired using WGS data from the UKBB Research Analysis Platform (https://www.ukbiobank.ac.uk/). Phenotypic data were derived from multiple sources, including International Classification of Diseases, 10th Revision (ICD-10) diagnoses (field 41270), PD status (field 131023), genetic ancestry grouping (field 22006), and age at recruitment (field 21022). PD cases were defined based on ICD-10 codes or self-reported PD status. Proxy cases were defined as individuals with a parent or sibling affected by PD. Controls were restricted to participants without neurological or nervous system disorders (category 2406), no parental history of PD or dementia (fields 20107 and 20110), and no ICD-10–defined neurodegenerative conditions, including dementia, vascular dementia, frontotemporal dementia, amyotrophic lateral sclerosis, parkinsonism, PD, progressive supranuclear palsy, or multiple system atrophy. McGill University Research Ethics Board granted ethics approval for this study.

### 2.2 Sequencing and quality control for PGK1

Variants within the *PGK1* locus (chrX:77,910,739–78,126,949; GRCh38) were extracted together with variants located within a ±100 kb flanking region upstream and downstream of the gene for common variants analyses. WGS data from the AMP-PD cohorts underwent quality control (QC) at both the individual and variant levels, following protocols described by (https://amp-pd.org/whole-genome-data) (Iwaki et al. 2021). For the UKBB WGS data, we conducted QC using the Genome Analysis Toolkit (GATK, v3.8), applying a minimum sequencing depth of 30x and a genotyping quality (GQ) threshold of 20 for inclusion in the analysis. Both AMP-PD and UKBB datasets were aligned to the human reference genome GRCh38. We used ANNOVAR to functionally annotate genomic variations (Yang and Wang 2015).

### 2.3 Statistical analysis

To account for differences in allele dosage of this X-linked gene, we performed analysis stratifying cohorts by sex and then combined. All results were meta-analyzed across cohorts. We assessed the associations between common variants (minor allele frequency MAF > 0.01) with PD, using logistic regression adjusted for age and principal components (PC) 1-5, implemented in PLINK v1.9 (Purcell et al. 2007). Bonferroni correction for multiple testing was applied. For rare variant analysis (MAF < 0.01) only variants within *PGK1*, we used the optimized sequence Kernel association (SKAT-O) test to assess the association with PD adjusting for age and PC1-5. To address the large number of UKBB controls and minimize bias due to case-control imbalance, we randomly selected a subset of controls at a 1:10 case to control ratio for both the cases only and cases plus proxies datasets. These subsets were subsequently used in rare variant analyses, and the results were combined using meta-analysis. We examined the burden of five variant groups in each cohort i) all rare, ii) functional (involving splicing, stop/frameshift, and nonsynonymous variants),iii) Combined Annotation Dependent Deletion (CADD) score ≥20, iv) nonsynonymous and v) loss-of-function variants. We applied Benjamini-Hochberg false discovery rate (FDR) method for multiple testing correction and performed a meta-analysis using metaSKAT package (Lee et al. 2013).

## Results

We identified 213 and 387 common variants (MAF>1%) in AMP-PD and UKBB, respectively. None was significantly associated with PD after Bonferroni correction (Table S1). For rare variant analyses (MAF<1%), we identified 357 rare variants in AMP-PD, including 1 high-CADD, 12 functional, 11 non-synonymous and 1 loss-of-function. UKBB cohort had 688 rare variants, including 5 high-CADD, 13 functional, 13 nonsynonymous (Table S2). SKAT-O analysis showed the same nominal association for both functional and nonsynonymous variants in sex-stratified analyses of UKBB males when proxy cases were included (P = 0.0475 for both). However, neither association remained significant after FDR correction (Pfdr = 1.00 and 0.6408, respectively; Table S3). In addition, no associations remained significant after meta-analysis across combined or sex-stratified cohorts for any variant category. The loss-of-function variant, p.G254Sfs*18 (c.758_759insCTCGG), was observed exclusively in PD cases (3 cases, 0 controls). This variant is a frameshift insertion in exon 8 that is predicted to introduce a premature termination codon. The three carriers were hemizygous males originating from AMP-PD, with ages at onset in the 60-70 years ranges. This variant has zero allele frequency reported on gnomAD and no reported significance.

We then examined four previously reported *PGK1* pathogenic variants associated with PGK1 deficiency and parkinsonism (Morales-Briceño et al. 2019; Virmani et al. 2014; Baba et al. 2018; Guimarães et al. 2024). None of these variants were identified in AMP-PD or UKBB (Table 2), nor reported in the GP2 browser (https://gp2.broadinstitute.org/) or gnomAD (https://gnomad.broadinstitute.org/).

**Table 2.** Reported *PGK1* pathogenic variants associated with parkinsonism, clinical features and evaluation in studied cohorts.

| Variant | Nucleotide Change | Phenotype with Parkinsonism | Levodopa Response | N patients | Reference(s) |
| --- | --- | --- | --- | --- | --- |
| p.Asp164Val | c.491A>T | Hemolytic anemia, epilepsy, stroke-like episodes, early-onset parkinsonism | Yes | 4 patients | (Morales-Briceño et al. 2019)<br>(Echaniz-Laguna et al. 2019) |
| p.Gly317Asp | c.950G>A | Cognitive impairment, epilepsy, psychiatric symptoms, resting/postural/kinetic tremor, bradykinesia, dystonia | Yes | 3 patients | (Guimarães et al. 2024) |
| p.Ala354Pro | c.1060G>C | Early-onset Parkinsonism (hemizygous); and adult-onset parkinsonism (heterozygous carrier) | Partial | 2 individuals, including a heterozygous carrier; 1 individual with dystonia/leuko dystrophy | (Morimoto et al. 2003);<br>(Sakaue et al. 2017)<br>(Baba et al. 2018) |
| p.Thr378Pro | c.1132A>C | Myopathy, exercise-induced myoglobinuria, developmental delay, progressive early onset parkinsonism (tremor, rigidity, bradykinesia, dysphagia) | Yes | 2 male patients | (Sotiriou et al. 2010);<br>(Virmani et al. 2014)<br>2026-08-18 15:43:00 |

Key: N, Number

## Discussion

In this study, we investigated the association between common and rare variants in *PGK1* and PD across two cohorts. No significant associations were observed after multiple correction or meta-analysis. These results suggest that *PGK1* variants are not associated with PD risk in the studied cohorts.

Although previous studies have linked PGK1 deficiency to parkinsonism (Morales-Briceño et al. 2019; Guimarães et al. 2024), to the best of our knowledge, a genetic association between *PGK1* and PD has not been studied. Nonetheless, *PGK1* has been investigated as a therapeutic target in PD. Pharmacoepidemiologic studies and a recent meta-analysis have reported an association between the use of glycolysis-enhancing alpha −1 blockers and reduced risk of developing PD (Ribeiro et al. 2024), though these findings await confirmation in randomized controlled trials.

This study has several limitations. The identified *PGK1* loss-of-function variant in AMP-PD cohort was not manually inspected using the Integrative Genomics Viewer to confirm the underlying read alignments. Although the variant passed the applied QC, manual review of sequencing reads can help distinguish true variants from sequencing or alignment artifacts, particularly for rare frameshift insertions and deletions. Therefore, the presence of this variant should be interpreted with caution. Secondly, only two cohorts were included, resulting in a relatively small sample size and reduced statistical power particularly for rare variant and sex-stratified analyses. Thirdly, our cohorts predominantly consisted of individuals of European ancestry, which limited the genetic diversity represented in the analysis. Finally, this study focused on PD risk and did not assess age at onset, disease progression, levodopa response, epilepsy, myopathy, hemolysis, metabolic biomarkers, or other phenotypes that may be more directly relevant to PGK1 deficiency biology. Therefore, absence of genetic association does not exclude a role for *PGK1* in PD pathophysiology or treatment response. PGK1 may still be relevant through altered glycolysis, neuronal energy metabolism, or pharmacological modulation and may lead to parkinsonism, however, our data does not support *PGK1* variation as a major independent genetic risk factor for PD.

In conclusion, we did not find evidence supporting association between *PGK1* and PD in these cohorts. However, very rare pathogenic *PGK1* variants may still contribute to syndromic parkinsonism. Future research could explore the loss-of-function variant, p.G254Sfs*18 in larger datasets, incorporate detailed phenotyping and potential interactions between *PGK1* and other PD-related genes.

## Author Roles

(1) Research Project: A. Conception, B. Organization, C. Execution; (2) Statistical Analysis: A. Design, B. Execution, C. Review and Critique; (3) Manuscript Preparation: A. Writing of the First Draft, B. Review and Critique.

L.V.C.: 1A, 1B, 1C, 2A, 2B, 3A. S.C.P.: 2B, 3B.

L.L.: 2B, 3B.

E.Y.: 2B.3B.

Z.G.O.: 1A, 1B, 2A, 3B.

K.S.: 1A,1B, 2A, 3B.

## Acknowledgements

We would like to sincerely thank the participants from the various cohorts who contributed to this study. This research was partially funded by the Canada First Research Excellence Fund through McGill University’s Healthy Brains, Healthy Lives initiative, with additional support from Calcul Quebec and Compute Canada. ZGO is supported by the Chercheurs-boursiers award from the Fonds de recherche du Québec – Santé (FRQS) and is a William Dawson Scholar. Access to UK Biobank data was enabled by the NeuroHub infrastructure and was undertaken thanks in part to funding from the Canada First Research Excellence Fund, awarded through the Healthy Brains, Healthy Lives initiative at McGill University. This research has been conducted using the UK Biobank Resource under Application Number 45551. Furthermore, data used in this article was also obtained on 10 June 2025 from the Accelerating Medicines Partnership® (AMP®) Parkinson’s Disease (AMP PD) Knowledge Platform, release version 4.2. The AMP® PD program is a public-private partnership managed by the Foundation for the National Institutes of Health and funded by the National Institute of Neurological Disorders and Stroke (NINDS) in partnership with the Food and Drug Administration (FDA), National Institute on Aging (NIA), Aligning Science Across Parkinson’s (ASAP) initiative; Celgene Corporation, a subsidiary of Bristol-Myers Squibb Company; GlaxoSmithKline plc (GSK); The Michael J. Fox Foundation for Parkinson’s Research (MJFF); AbbVie Inc.; Pfizer Inc.; Sanofi US Services Inc.; and Verily Life Sciences LLC.

The Harvard Biomarker Study (HBS) is a collaboration of HBS investigators and funded through philanthropy and NIH and Non-NIH funding sources. The Stephen & Denise Adams Center for Parkinson’s Disease Research of Yale School of Medicine is funded through philanthropy and NIH and non-NIH funding sources. The HBS and CPDR-Y Investigators have not participated in reviewing the data analysis or content of the manuscript.

BioFIND is sponsored by The Michael J. Fox Foundation for Parkinson’s Research (MJFF) with support from the National Institute for Neurological Disorders and Stroke (NINDS). The BioFIND Investigators have not participated in reviewing the data analysis or content of the manuscript.

The Parkinson’s Disease Biomarker Program (PDBP) consortium is supported by the National Institute of Neurological Disorders and Stroke (NINDS) at the National Institutes of Health. A full list of PDBP investigators can be found at https://pdbp.ninds.nih.gov/policy. The PDBP investigators have not participated in reviewing the data analysis or content of the manuscript.

The Study of Isradipine as a Disease-modifying Agent in Subjects With Early Parkinson Disease, Phase 3 (STEADY-PD3) is funded by the National Institute of Neurological Disorders and Stroke (NINDS) at the National Institutes of Health with support from The Michael J. Fox Foundation and the Parkinson Study Group. The STEADY-PD3 investigators have not participated in reviewing the data analysis or content of the manuscript.

Genome sequence data for the Lewy body dementia case-control cohort were generated at the Intramural Research Program of the U.S. National Institutes of Health. The study was supported in part by the National Institute on Aging (program #: 1ZIAAG000935) and the National Institute of Neurological Disorders and Stroke (program #: 1ZIANS003154).

## Data availability

All the data generated in this study are provided within the manuscript. https://github.com/gan-orlab/PGK1-on-PD

